## Appendix S1 for "The influence of cross-border mobility on the COVID-19 epidemic in Nordic countries"

November 2, 2023

Shubin Mikhail[1 *], Hilde Kjelgaard Brustad[2], Jørgen Eriksson Midtbø[3], Felix Günther[4], Laura Alessandretti[5], Tapio Ala-Nissila[6, 7], Gianpaolo Scalia Tomba[4, 8], Mikko Kivelä[9], Louis Yat Hin Chan[3], Lasse Leskelä[1]

**1** Department of Mathematics and Systems Analysis, Aalto University, Espoo, Finland
**2** Oslo Center for Biostatistics and Epidemiology, Oslo University Hospital, Norway
**3** Department of Method Development and Analytics, Norwegian Institute of Public Health, Oslo, Norway
**4** Department of Mathematics, Stockholm University, Sweden
**5** DTU Compute, Technical University of Denmark, Copenhagen, Denmark
**6** Quantum Technology Finland Center of Excellence, Department of Applied Physics, Aalto University
**7** Interdisciplinary Centre for Mathematical Modelling and Department of Mathematical Sciences, Loughborough University, United Kingdom
**8** Department of Mathematics, University of Rome Tor Vergata, Italy
**9** Department of Computer Science, Aalto University, Espoo, Finland

*

### Mobility data

In this Appendix we give a detailed description of the different sources of the mobility data and how the data are aggregated.

The mobility data consist of road traffic data, airliner data, rail data and ferry traffic data. The different data sources are combined for each combination of countries (Norway, Sweden, Denmark, Finland and outside), but some transportation modes, or the corresponding data, are not available for certain combinations of countries. Table 1 lists the data sources available for the different combinations of countries, rows specifying the source country of travel and columns the destination country.

| Source \ Destination | DEN | FIN | NOR | SWE | Abroad |
|---|---|---|---|---|---|
| DEN | | X<br>Air<br>X<br>X | X<br>Air<br>Ferry<br>X | Road<br>Air<br>Ferry<br>Rail | Road<br>e<br>Ferry<br>Rail |
| FIN | X<br>Air<br>X<br>X | | Road<br>Air<br>Ferry<br>X | Road<br>Air<br>Ferry<br>X | Road<br>e<br>Ferry<br>Rail |
| NOR | X<br>Air<br>Ferry<br>X | Road<br>Air<br>Ferry<br>X | | Road<br>Air<br>Ferry<br>Rail | Road<br>e<br>Ferry<br>X |
| SWE | Road<br>Air<br>Ferry<br>Rail | Road<br>Air<br>Ferry<br>X | Road<br>Air<br>Ferry<br>Rail | | X<br>e<br>Ferry<br>X |
| Abroad/outside | Road<br>Air<br>Ferry<br>Rail | Road<br>Air<br>Ferry<br>Rail | Road<br>Air<br>Ferry<br>X | X<br>Air<br>Ferry<br>X | |

Table 1: A table of all available data sources (road, air, ferry and rail, as shown in the cells) for the different combinations of Nordic countries and outside. The symbol e denotes data that we have estimated from the available information while X denotes irrelevant or non-existent data source for the specific combination of countries. The rows show source country while the columns show the country of destination.

Travel restrictions during the pandemic depended on the purpose of the travel. For instance, commuters from neighbouring countries were allowed to enter Norway at times where the borders were closed. The road traffic data are therefore divided into long-term travellers and short-term travellers (daily commuters). Road-traffic data are the only data source with the appropriate time resolution for separating commuting from long-term travel. However, commuting between Sweden and Denmark also happens by railway through the Øresund-Öresund bridge, which needs to be taken into account for short-term travel flows.

The number of individuals travelling from source country $s$ to destination country $d$ on day $t$, for $s, d \in \{\mathrm{DEN}, \mathrm{FIN}, \mathrm{NOR}, \mathrm{SWE}\}$ *i.e.*, between the Nordic countries, are denoted $D^{\mathrm{long}}_{t,d\leftarrow s}$ and $D^{\mathrm{short}}_{t,d\leftarrow s}$ for long-term and short-term travellers, respectively. We further denote the number of individuals, traveling long or short term, from abroad/outside to the Nordic countries by $D^{\mathrm{long}}_{t,d\leftarrow o}$ and $D^{\mathrm{short}}_{t,d\leftarrow o}$ for $d \in \{\mathrm{DEN}, \mathrm{FIN}, \mathrm{NOR}, \mathrm{SWE}\}$. Correspondingly, the number of individuals leaving the Nordic countries, who traveled either long or short-term, are denoted $D^{\mathrm{long}}_{t,o\leftarrow s}$ and $D^{\mathrm{short}}_{t,o\leftarrow s}$ for $s \in \{\mathrm{DEN}, \mathrm{FIN}, \mathrm{NOR}, \mathrm{SWE}\}$, respectively. Figure 1 in the main paper summarizes schematically the land connections between the four Nordic countries.

### Road traffic

For the inter-Nordic road traffic we collected data from sensors located at the borders of Norway, Sweden, Denmark and Finland. At each border crossing there are two sensors, separately recording traffic in both directions. The raw data give either the number of vehicles passing a sensor each hour or the exact time stamp of each vehicle passing the sensor. We aggregated the data such that data for each sensor specifies the number of vehicles passing the sensor during each hour. Road traffic data at 7 locations (14 sensors) along the Denmark Germany border were acquired from The Danish Road Directorate by request. Data for the border between Sweden and Denmark, across the Øresund-Öresund bridge, were acquired through contact with the Swedish Transport Administrations, who gather the data from Øresundsbro Konsortiet's API. The rest of the traffic data for the Swedish border were extracted from the Norwegian Public Roads Administration's Traffic Data API and Fintraffic's API Digitraffic. Data were gathered from 26 locations (52 sensors) along the Swedish-Norwegian border and from 5 locations (10 sensors) locations along the Swedish-Finnish border. Along the Norwegian-Finnish border, data from 6 locations (12 sensors) were extracted from the Norwegian API. Data for the Finnish-Russian border (4 locations, 8 sensors) and the Norwegian-Russian border (1 locations, 2 sensors) were extracted from the Finnish and Norwegian API, respectively.

We estimate the number of individuals travelling from one country in the morning and back to the opposite direction in the evening. These individuals count as the short-term travellers. We make the assumption that distribution of travellers throughout the day is generated by the weighted sum of different Gaussian distributions. Each of these distributions then corresponds to a specific type of traveller. For each sensor, we therefore fit a Gaussian Mixture Model (GMM) to the number of travellers, for each day of the week separately. Then, for each sensor and each day of the week, we have $K = 10$ sets of estimated parameters (mean, variance and a weight) specifying the fitted Gaussian components.

Next we need to identify what components correspond to commuting for each sensor. We look at the two sensors at the same location, with traffic in opposite directions. Let's call these sensors $s_1$ and $s_2$, where $s_1$ is the sensor of focus. If we find at least one component for sensor $s_1$ that has mean between 5.30 am and 9 am in the morning with weight higher or equal to 0.1, and at least one component for sensor $s_2$ with mean between 2 pm and 6 pm in the evening with weight higher or equal to 0.1, we label these components as the commuting components that correspond to commuting for sensor $s_1$.

We then compute the probability that each travel throughout the day is generated by either of these components, for sensor $s_1$ and $s_2$ separately. By summing this for each travel throughout the day, we get the estimated number of daily commuters in direction 1, $N^{\mathrm{short}}_1$ (mostly consisting of the morning wave of commuters passing sensor $s_1$) and the estimated number of daily commuters returning back in direction 2, $N^{\mathrm{short}}_1$ (mostly consisting of the evening wave of commuters passing sensor $s_2$). For some pairs of sensors, the total traffic flows in each directions do not cancel out, which causes the morning and evening flows of commuters to be unbalanced. We therefore decide that the daily number of estimated commuters for sensor $s_1$ is $N^{\mathrm{short}}_1 = \min(N^{\mathrm{short}}_1, N^{\mathrm{short}}_2)$. The same process is then performed again with $s_2$ as the sensor in focus.

For each pair of countries, the daily number of estimated commuters are summed over the sensors in each direction, resulting in two time series:

$$D^{\mathrm{all,road}}_{t,d\leftarrow s} \quad \text{and} \quad D^{\mathrm{short,road}}_{t,d\leftarrow s},$$

for a combination of source country $s$ and destination country $d$, where road traffic data are available. The superscript *all* refers to the total number of vehicles before estimating the commuters. We make the assumption that commuters travel alone in the vehicle, meaning that the number of commuter vehicles equals the number of individuals. For the long-term travelers, we multiply the number of vehicles by a factor of 1.2 to account for passengers in the vehicles. The number of long-term travelers by road from country $s$ to country $d$ is then given by

$$D^{\mathrm{long,road}}_{t,d\leftarrow s} = 1.2 \times \left(D^{\mathrm{all,road}}_{t,d\leftarrow s} - D^{\mathrm{short,road}}_{t,d\leftarrow s} - D^{\mathrm{short,road}}_{t,d\rightarrow s}\right).$$

Figures 1, 2 and 3 show the the daily number of vehicles, as a 7-day moving average, for all combinations of countries where road traffic if possible, for short-term travel (green line), long-term travel (red line) and total number of vehicles (blue line). The right and left plot in each row show the travel in each direction for a pair of countries.

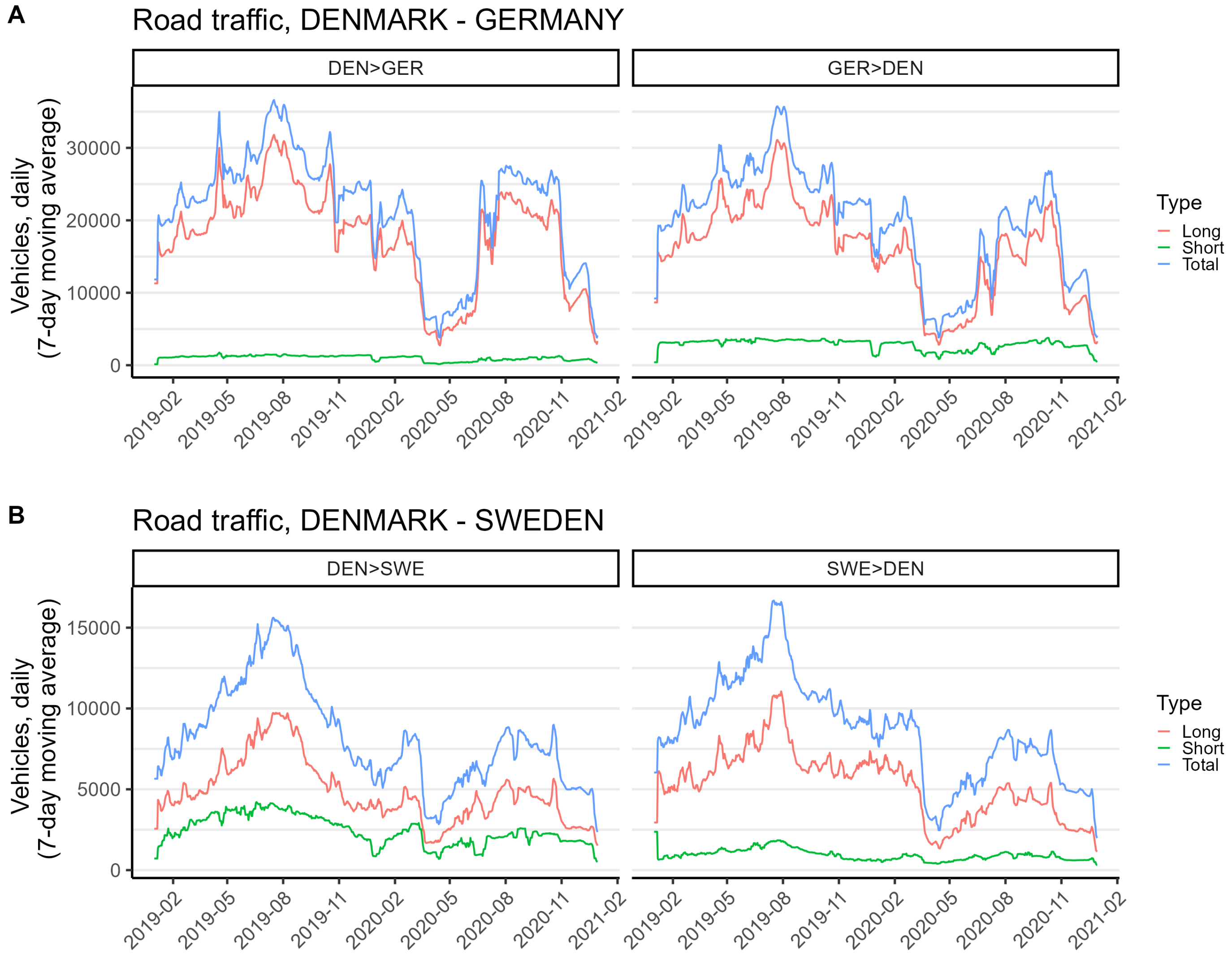


Figure 1: Daily number of vehicles in 2019 and 2020, as a 7-day moving average for travel between (A) Denmark and Germany and (B) Denmark and Sweden, divided into short-term travel (green line), long-term travel (red line) and total number of vehicles (blue line).

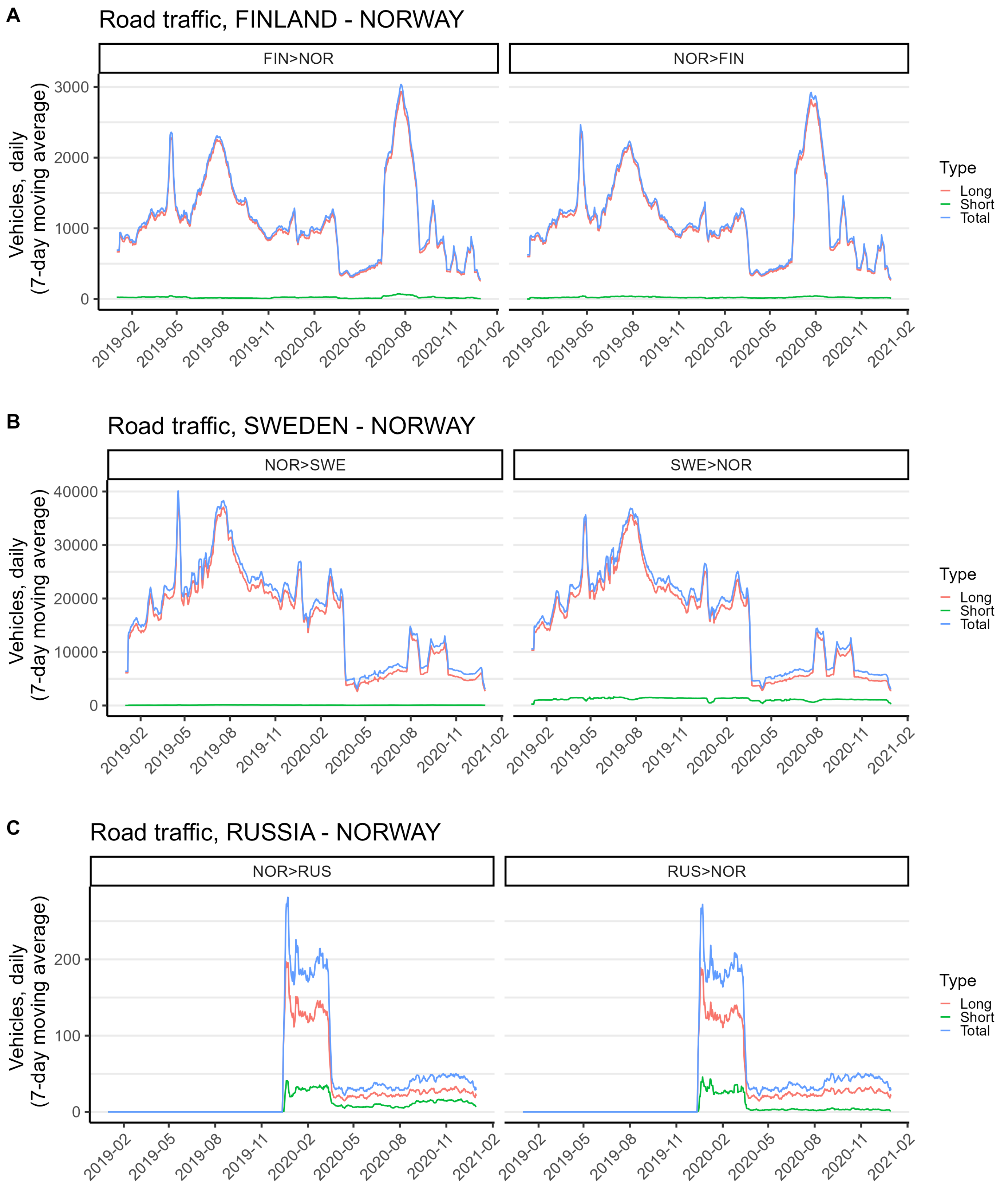


Figure 2: Daily number of vehicles in 2019 and 2020, as a 7-day moving average for travel between (A) Finland and Norway, (B) Sweden and Norway and (C) Russia and Norway, divided into short-term travel (green line), long-term travel (red line) and total number of vehicles (blue line).

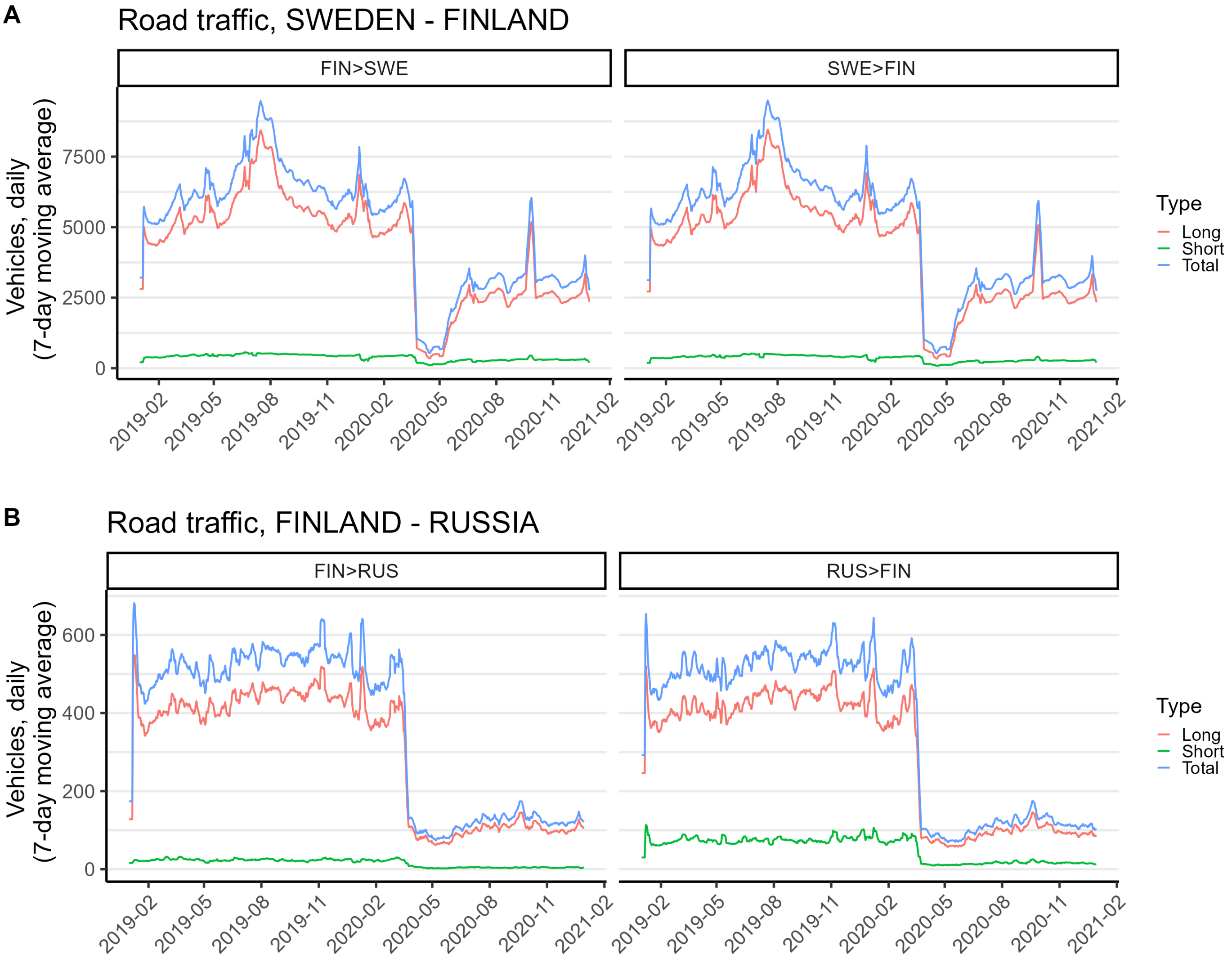


Figure 3: Daily number of vehicles in 2019 and 2020, as a 7-day moving average for travel between (A) Sweden and Finland and (B) Finland and Russia, divided into short term travel (green line), long term travel (red line) and total number of vehicles (blue line).

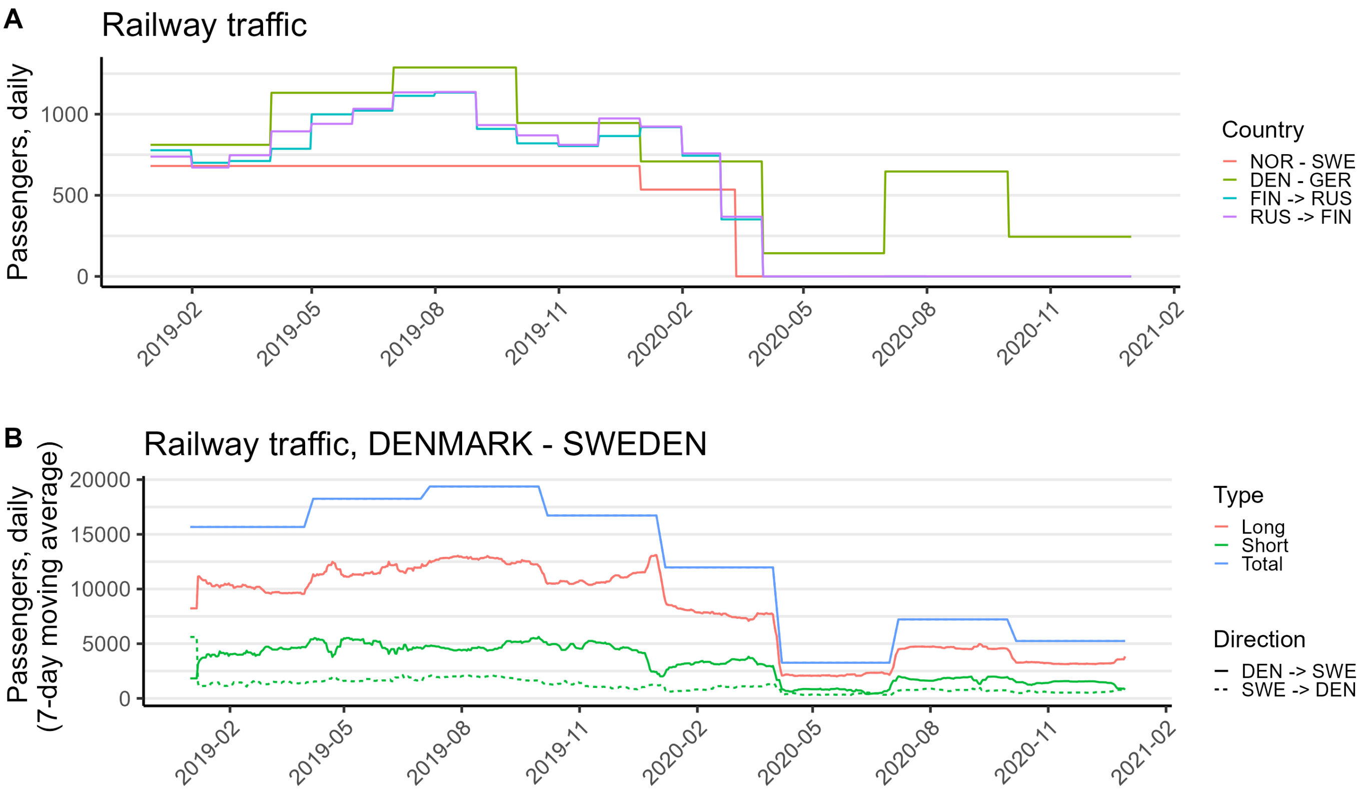


Figure 4: (A) Daily number of railway passengers in 2019 and 2020 between Norway and Sweden, Denmark and Germany, and Finland and Russia. (B) Daily number of railway passengers in 2019 and 2020, as a 7-day moving average, between Denmark and Sweden, divided into long-term travelers (red line), short-term travelers (green line) and total number of travelers (blue line). The solid lines denote travelers from Denmark to Sweden, while the dashed line shows travelers from Sweden to Denmark.

### Railway traffic

The annual number of railway passengers between Norway and Sweden were acquired from the report "Jernbanestatistikk-2020" published by the Norwegian Railway Directorate. The total number of passengers crossing the border in both directions by train was 497,000 during 2019. For the year 2020 the number of passengers was 76,000. For 2019 we distribute the passengers evenly in both directions and for each day throughout the year, resulting in 681 daily travelers in each direction. When lockdown started in Norway in the middle of March 2020, railway traffic between Norway and Sweden was drastically reduced. We therefore distribute the railway passengers for 2020 evenly in both directions from the beginning of 2020 until the lockdown started. This means 535 passengers daily in each direction between January 1, 2020 and March 11, 2020. For the rest of 2020, we assume no railway traffic between Norway and Sweden.

Railway traffic data to and from Denmark were acquired from Statistics Denmark, table BANE25. This is quarterly statistics on the number of railway passengers between Denmark and Sweden, and between Denmark and Germany. For each pair of countries, we distributed the number of passengers evenly in both directions and across each day of the three-month periods.

There is commuting between Denmark and Sweden by train across the Øresund-Öresund Bridge. The quarterly railway statistics do not have high enough time resolution to apply the same method for estimating short-term travellers as for the road traffic data. As an alternative method, we looked at the daily fraction of short-term travellers among all travelers for the road traffic data, $r^{\text{short,road}}_{t,d\leftarrow s}$, and assumed this fraction of daily railway passengers between Denmark and Sweden to be short term travellers:

$$D^{\text{short,railway}}_{t,d\leftarrow s} = r^{\text{short,road}}_{t,d\leftarrow s} D^{\text{all,railway}}_{t,d\leftarrow s},$$

for $s, d \in \{\text{SWE}, \text{DEN}\}$. The daily number of long-term travellers by train between Sweden and Denmark are then computed in the same way as for the road traffic data:

$$D^{\text{long,railway}}_{t,d\leftarrow s} = D^{\text{all,railway}}_{t,d\leftarrow s} - D^{\text{short,railway}}_{t,d\leftarrow s} - D^{\text{short,railway}}_{t,d\rightarrow s}.$$

The last possibility of railway traffic is between Finland and Russia. There are mandatory border controls between the countries and the Finnish customs provide tables of traffic volumes at Finnish borders. This includes monthly statistics on the number of railway passengers for each direction separately.

Figure 4A shows the daily number of railway passengers in 2019 and 2020 between Norway and Sweden, Denmark and Germany, and Finland and Russia. For traffic between Norway and Sweden, and Denmark and Germany, only one time series for each of the two combinations of countries is needed, because the traffic flow is assumed to be equal in both directions. Finland and Russia, on the other hand, have directed traffic flows. Figure 4B shows the daily number of railway passengers, as a 7-day moving average, between Denmark and Sweden, divided into long-term travelers (red line), short-term travelers (green line), and total number of travelers (blue line). The solid lines denote travelers from Denmark to Sweden, while the dashed line shows travelers from Sweden to Denmark. Note that the because the total number of travelers are equal in both directions due to lack of directed traffic flows in the raw data (blue line), the number of long-term travelers is the same in both directions even though that of the short-term travelers differ.

For convenince, we have collected the relevant links for the data below:

1. NOR-SWE: `https://www.jernbanedirektoratet.no/contentassets/e71b740c9f5d4583aed0c193c11faec7/jernbanes pdf`.
2. GER-DEN: `https://www.statbank.dk`, Table BANE25.
3. FIN-RUS: `https://tulli.fi/en/statistics/statistics-on-logistics`

### Air traffic

Air traffic data are provided by European Commission Joint Research Centre (JRC). The data are downscaled from monthly statistics. The data give the daily number of air passengers with destination to Norway, Sweden, Denmark or Finland, *i.e.*, travel between the Nordic countries and travel from abroad to Nordic countries, stratified by a non-Nordic source country. Data on individuals leaving the Nordic countries are missing in this data source. We therefore balance the inflow and outflow of the Nordic countries by assuming the same number of travellers in both directions, *i.e.*,

$$D^{\text{air,long}}_{t,o\leftarrow d} = D^{\text{air,long}}_{t,d\leftarrow o}$$

for $d \in \{\text{DEN}, \text{FIN}, \text{NOR}, \text{SWE}\}$. The air traffic data only contribute to long-term travelling. The raw data are not publicly available.

### Ferry traffic

Ferry traffic data contain the number of ferry passengers travelling to and from the Nordic countries and abroad. Data were acquired from Statistics Norway, Transport Analysis, Sweden, Statistics Denmark and Statistics Finland. The data from Statistics Norway and Transport Analysis, Sweden are on a quarterly time scale, while data from Statistics Finland and Statistics Denmark have a monthly temporal resolution. The number of passengers are downscaled to a daily resolution by distributing them evenly for each day of the time period. Data from Statistics Denmark only specify the total number of travelers in both directions, and we distribute these evenly in both directions. Some combinations of countries are contained in data sets from two sources, *e.g.*, traffic between Norway and Denmark is contained both in the data set from Statistics Denmark and Statistics Norway. The source with the highest temporal resolution (monthly) is then selected. We note that there is considerable amount of ferry traffic between Finland and Estonia, and these passengers were put in the long-term category.

The following links contain the relevant data sources for ferry traffic:

1. FIN: `https://pxdata.stat.fi/PxWeb/pxweb/en/StatFin/` "Table 12j3 – Passenger Traffic between Finland and foreign countries by port and country, monthly, 2016M01-2022M09".
2. NOR: `https://www.ssb.no/en/statbank/` "Table 04225: Ferry transport between Norway and foreign countries. Number of units, by port and relation, direction and type of cargo 2003K1 - 2022K2".
3. SWE: `https://www.trafa.se/en/maritime-transport/shipping-goods/` (not used).
4. DEN: `https://statbank.dk` Table SKIB34.

### Combining all data sources

All the available data sources (road, train, ferry and air) contribute to long-term travelers. Air traffic data and ferry traffic data contribute to the long-term travelers only. The total number of long-term travellers between the Nordic countries is given by

$$D^{\text{long}}_{t,d\leftarrow s} = D^{\text{long,road}}_{t,d\leftarrow s} + D^{\text{long,railway}}_{t,d\leftarrow s} + D^{\text{long,ferry}}_{t,d\leftarrow s} + D^{\text{long,air}}_{t,d\leftarrow s}$$

for $s, d \in \{\text{DEN}, \text{FIN}, \text{NOR}, \text{SWE}\}$.

Road traffic contributes to short-term travel between all the countries, while ferry traffic contributes to short-term travels between Sweden and Denmark only. Short-term travellers between the Nordic countries are given by

$$D^{\text{short}}_{t,d\leftarrow s} = D^{\text{short,road}}_{t,d\leftarrow s} + D^{\text{short,railway}}_{t,d\leftarrow s}, \quad D^{\text{short,railway}}_{t,d\leftarrow s} = 0 \text{ for } s, d \neq \{\text{SWE}, \text{DEN}\},$$

for $s, d \in \{\text{DEN}, \text{FIN}, \text{NOR}, \text{SWE}\}$. Obviously, some of these terms are identically zero unless each pair of countries is connected by a road or a trainline.

Travelers from outside of the four Nordics are added up to form a single time series of travellers from abroad to each of the Nordic countries, denoted by $D^{\text{long}}_{t,d\leftarrow o}$ and $\text{D}^{\text{short}}_{t,d\leftarrow o}$ for long-term and short-term travellers entering the Nordic countries, respectively, and $D^{\text{long}}_{t,o\leftarrow s}$ and $D^{\text{short}}_{t,o\leftarrow s}$ for travelers leaving the Nordic countries.

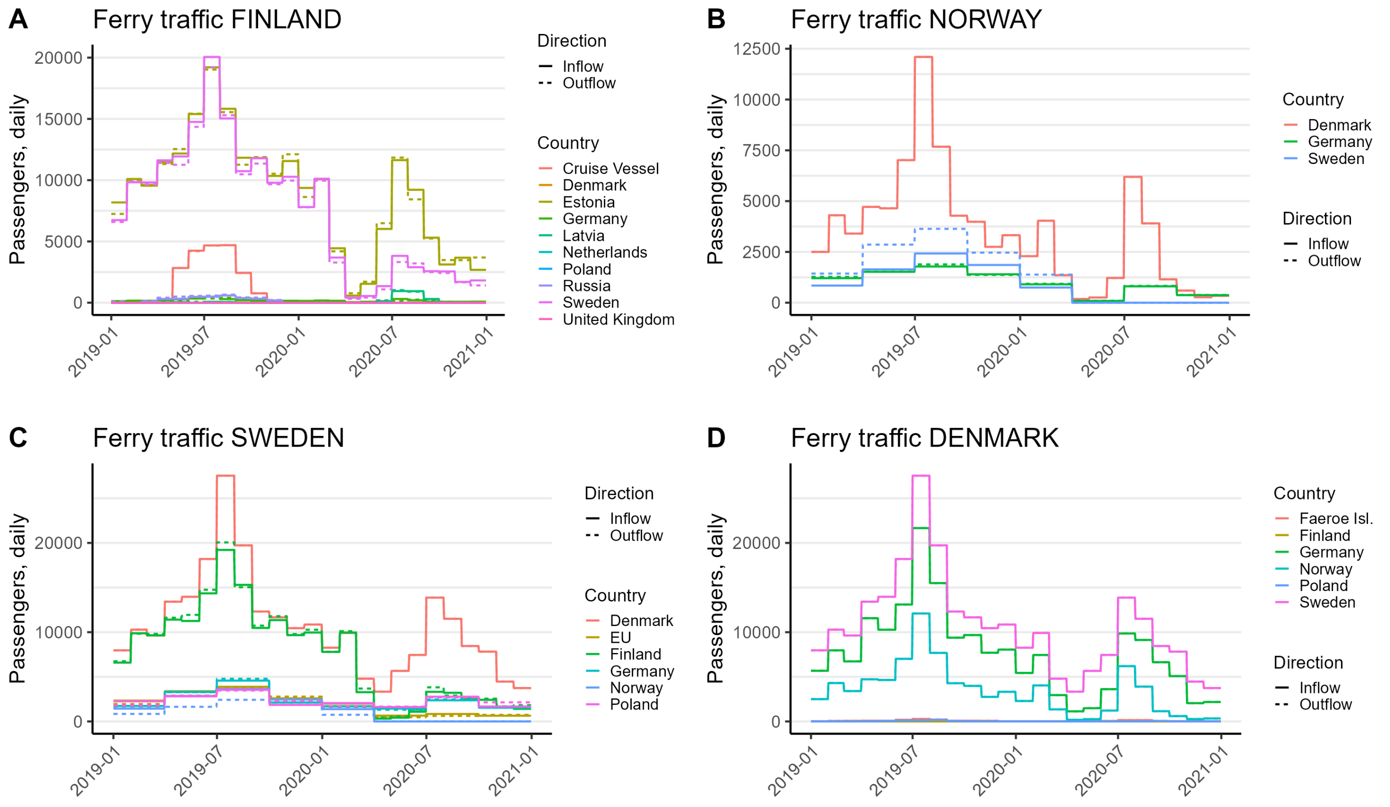


Figure 5: Daily number of ferry passengers arriving and departing in (A) Finland, (B) Norway, (C) Sweden and (D) Denmark. The different colours denote different countries. Solid lines denote inflow of traffic and dashed lines denote outflow of traffic.

### Estimated number of non-Nordic infected travellers entering the Nordic countries

The estimates of infected individuals entering a Nordic country from a non-Nordic country during the study period are based on COVID-19 incidence data from Our World in Data [1], `https://github.com/owid/covid-19-data/blob/master/public/data/owid-covid-data.csv` downloaded August 30, 2022. Among the available columns in this file, our estimates are based on the column "new_cases_smoothed_per_million", which is country and date specific. The reported numbers are converted to prevalence of individuals using the following procedure.

First, we estimate number of infections $I_{x,t}$ in the four modelled Nordic countries. We do it by fitting our model assuming there are no inflow from the non-modelled countries. We take a posterior mean as an estimate for $I_{x,t}$.

Second, knowing the true number of infections $I_{x,t}$ and the reported number of cases $C_{x,t}$ (taken from [1]) we can compute the reporting factor: number of reported cases per one actual infection $f_{x,t} = C_{x,t}/I_{x,t}$. See the Figure 6(A). The number of reported cases may be zero or missing. This is especially true for the beginning of the epidemic, when the surveillance systems were not established. To avoid extreme values of $f_{x,t}$, we apply a layer of Gaussian smoothing onto $C_x$. See the Figure 6(B). We then compute the final reporting factor $f_t$ by averaging out the values between the four Nordic countries.

Applying $f_t$ to the data in the above-mentioned file, the expected number of imported infections from a non-Nordic country $c$ to Nordic country $x$ on day $t$ is calculated as

$$i^{\text{inflow}}_{t,x\leftarrow c} = D^{\text{long}}_{t,x\leftarrow c} \times p_{t,c} \times 10^{-6}/f_t,$$

where $D^{\text{long}}_{t,x\leftarrow c}$ is the number of long-term travellers from a non-Nordic country $c$ to a Nordic country $x$ and $p_{t,c}$ is the prevalence $i_{t,c}$ expressed per one million inhabitants.

Finally, the contributions from each non-Nordic country is summed to yield the total estimate of infections imported from abroad on day $t$ to a Nordic country $x$, *i.e.*,

$$i^{\text{inflow}}_{t,x} = \sum_{c\in\text{non-Nordic}} i^{\text{inflow}}_{t,x\leftarrow c} = \sum_{c\in\text{non-Nordic}} \left(D^{\text{long}}_{t,x\leftarrow c} \times p_{t,c} \times 10^{-6}\right)/f_t.$$

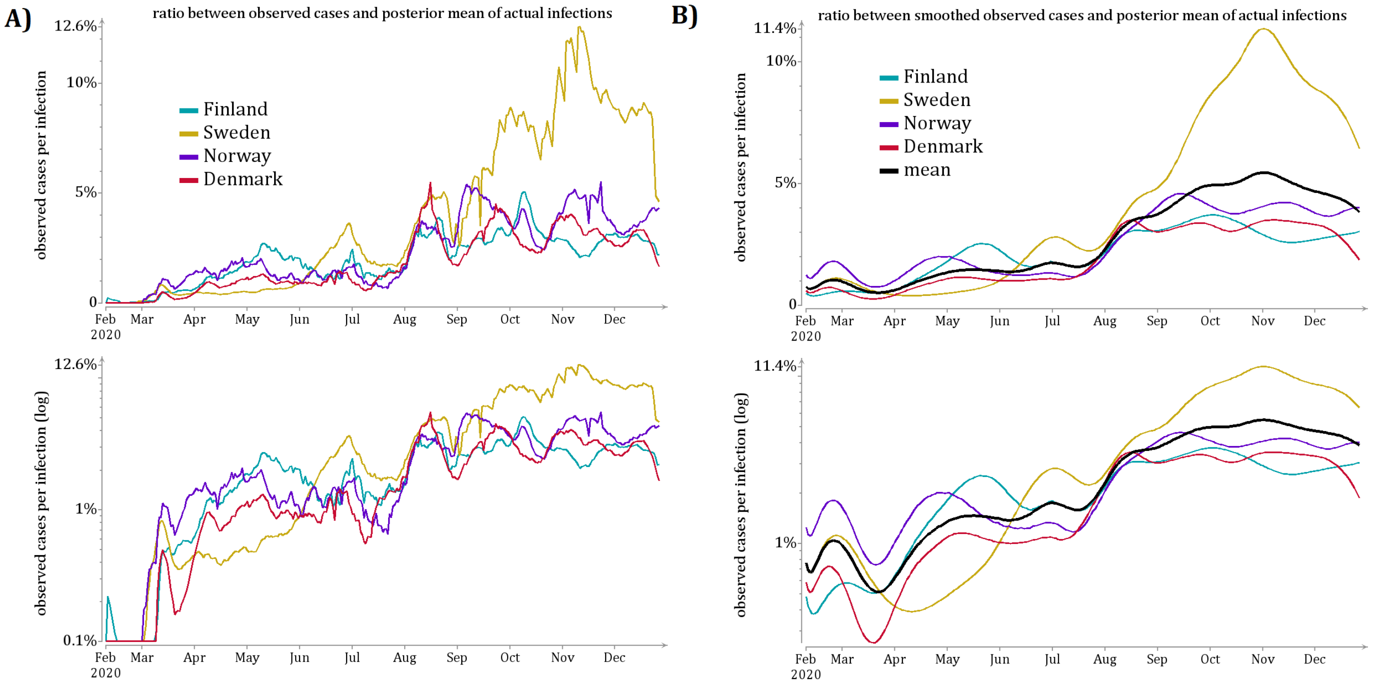


Figure 6: (A)the reporting factor: number of reported cases per one actual infection $f_x = C_x/I_x$ (B) same quantity after smoothing $C_x$.
