## Appendix S2 for "The influence of cross-border mobility on the COVID-19 epidemic in Nordic countries"

### The influence of cross-border mobility on the COVID-19 epidemic in Nordic countries
### Appendix S2: Derivation of the transmission model

November 2, 2023

Shubin Mikhail[1 *], Hilde Kjelgaard Brustad[2], Jørgen Eriksson Midtbø[3], Felix Günther[4], Laura Alessandretti[5], Tapio Ala-Nissila[6, 7], Gianpaolo Scalia Tomba[4, 8], Mikko Kivelä[9], Louis Yat Hin Chan[3], Lasse Leskelä[1]

**1** Department of Mathematics and Systems Analysis, Aalto University, Espoo, Finland
**2** Oslo Center for Biostatistics and Epidemiology, Oslo University Hospital, Norway
**3** Department of Method Development and Analytics, Norwegian Institute of Public Health, Oslo, Norway
**4** Department of Mathematics, Stockholm University, Sweden
**5** DTU Compute, Technical University of Denmark, Copenhagen, Denmark
**6** Quantum Technology Finland Center of Excellence, Department of Applied Physics, Aalto University
**7** Interdisciplinary Centre for Mathematical Modelling and Department of Mathematical Sciences, Loughborough University, United Kingdom
**8** Department of Mathematics, University of Rome Tor Vergata, Italy
**9** Department of Computer Science, Aalto University, Espoo, Finland

*

In this Appendix, we describe how we derive the mathematical model used in our work.

#### Basic definitions

Let us start with a basic Susceptible-Infected-Recovered (SIR) model in discrete time without mobility. Let $S_{t,x}$, $I_{t,x}$, $R_{t,x}$ be the numbers of Susceptible, Infected and Removed individuals on day $t$ in the country $x$; let $N_x$ be the population size and $\gamma$ - the recovery rate. The model is updated as follows:

$$\begin{aligned}
S_{t+1,x} &= S_{t,x} - r_{t,x} S_{t,x};\\
I_{t+1,x} &= I_{t,x} + r_{t,x} S_{t,x} - \gamma I_{t,x};\\
R_{t+1,x} &= R_{t,x} + \gamma I_{t,x};\\
r_{t,x} &= \beta_{t,x} I_{t,x} / N_x.
\end{aligned}$$

Here $r_{t,x}$ is the risk of getting infected in country $x$ on day $t$ and $\beta_{t,x}$ is a country- and time-specific infectivity, defined as the average amount of secondary infections caused by a primary infection on day $t$ in country $x$, assuming everybody is susceptible.

In the model, effective reproduction number for country $x$ can be approximated by $R^{\text{eff}}_{t,x} \approx \beta_{t,x} \gamma^{-1} S_{t,x}/N_x$ Further, the population size $N_x = S_{t,x} + I_{t,x} + R_{t,x}$ remains constant in the model. The model can be visualized with the following flow diagram:

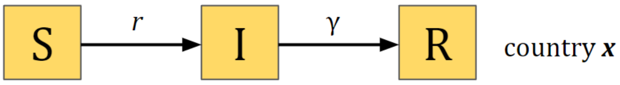


#### Adding long-term travellers between Nordic countries

Lets assume that mobility matrices $D^{\text{long}}_{t,d\leftarrow s}$ represent the number of individuals from the source country $s$ travelling to the destination country $d$ on day $t$ for a long-term visit. Let $p(S|t,d,s)$, $p(I|t,d,s)$ and $p(R|t,d,s)$ be the probability for these individuals to be Susceptible, Infectious, or Removed, respectively. The model is then updated as following:

$$
\begin{aligned}
S_{t+1,x} &= S_{t,x} - r_{t,x}S_{t,x} && + \sum_{s,s\neq x} D^{\text{long}}_{t,x\leftarrow s}\,p(S|t,x,s) && - \sum_{s,s\neq x} D^{\text{long}}_{t,d\leftarrow x}\,p(S|t,d,x)\\
I_{t+1,x} &= I_{t,c} + r_{t,x}S_{t,x} - \gamma I_{t,c} && + \sum_{s,s\neq x} D^{\text{long}}_{t,x\leftarrow s}\,p(I|t,x,s) && - \sum_{s,s\neq x} D^{\text{long}}_{t,d\leftarrow x}\,p(I|t,d,x)\\
R_{t+1,x} &= R_{t,c} + \gamma I_{t,c} && + \underbrace{\sum_{s,s\neq x} D^{\text{long}}_{t,x\leftarrow s}\,p(R|t,x,s)}_{\text{people arriving to country x}} && - \underbrace{\sum_{d,d\neq x} D^{\text{long}}_{t,d\leftarrow x}\,p(R|t,d,x)}_{\text{people leaving from country x}}
\end{aligned}
$$

In the simplest case, we assume that the probability of the traveller belonging to the certain compartment is equal to the prevalence of this compartment in the source country:

$$
\begin{aligned}
p(S|t,d,s) &= S_{t,s}/N_{t,s};\\
p(I|t,d,s) &= I_{t,s}/N_{t,s};\\
p(R|t,d,s) &= R_{t,s}/N_{t,s}.
\end{aligned}
$$

By defining a relative mobility matrix as

$$
M^{\text{long}}_{t,d\leftarrow s} = \begin{cases} \dfrac{D^{\text{long}}_{t,d\leftarrow s}}{N_{t,s}}, & \text{for } s \neq d \text{ (portion of population leaving to } d\text{);}\\[2ex] 1 - \sum_{x\neq s} \dfrac{D^{\text{long}}_{t,x\leftarrow s}}{N_{t,s}}, & \text{for } s = d \text{ (portion of population staying in } s\text{),} \end{cases}
$$

we can express the previous equations as

$$
\begin{aligned}
S_{t+1,x} &= S_{t,\bullet}M^{\text{long}}_{t,x\leftarrow\bullet} - r_{t,x}S_{t,x};\\
I_{t+1,x} &= I_{t,\bullet}M^{\text{long}}_{t,x\leftarrow\bullet} + r_{t,x}S_{t,x} - \gamma I_{t,c};\\
R_{t+1,x} &= R_{t,\bullet}M^{\text{long}}_{t,x\leftarrow\bullet} + \gamma I_{t,x};\\
r_{t,x} &= \beta_{t,x}I_{t,x}/N_{t,x}.
\end{aligned}
$$

In this model, the population sizes $N_{x,t}$ change with time as $N_{x,t+1} = N_{t,\bullet}M^{\text{long}}_{t,x\leftarrow\bullet}$. However, in the present case these changes are $\leq 3\%$. The total size of the population $\sum_x N_{t,x}$ remains constant. This is visualized in the graph below.

Changes in population size due to long-term travelling between Nordics

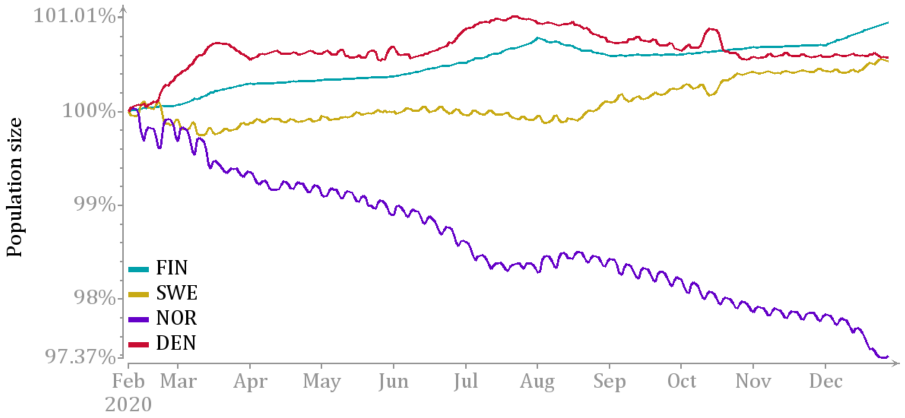


The model can be visualized with the following flow chart:

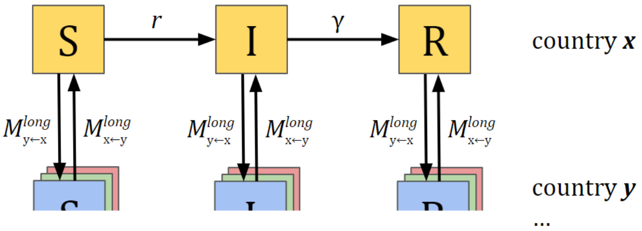

#### Adding long-term travellers between Nordic and other countries

We denote the rest of the world as a *country* $\circ$ (outside world). Let be $D^{\text{long}}_{t,d\leftarrow\circ}$ be the number of individuals arriving to the country $d$ from a non-Nordic country for a long-term visit, and $D^{\text{long}}_{t,\circ\leftarrow d}$ be the number of individuals leaving to the non-Nordic country $\circ$. We then update the model as follows:

$$\begin{aligned}
S_{t+1,x} &= S_{t,\bullet}M^{\text{long}}_{t,x\leftarrow\bullet} - r_{t,x}S_{t,x} && +D^{\text{long}}_{t,x\leftarrow\circ}p(S|t,x,\circ) && -D^{\text{long}}_{t,\circ\leftarrow x}p(S|t,\circ,x);\\
I_{t+1,x} &= I_{t,\bullet}M^{\text{long}}_{t,x\leftarrow\bullet} + r_{t,x}S_{t,x} - \gamma I_{t,c} && +D^{\text{long}}_{t,x\leftarrow\circ}p(I|t,x,\circ) && -D^{\text{long}}_{t,\circ\leftarrow x}p(I|t,\circ,x);\\
R_{t+1,x} &= R_{t,\bullet}M^{\text{long}}_{t,x\leftarrow\bullet} + \gamma I_{t,c} && \underbrace{+D^{\text{long}}_{t,x\leftarrow\circ}p(R|t,x,\circ)}_{\text{people arriving to country } x \text{ from outside Nordics}} && \underbrace{-D^{\text{long}}_{t,\circ\leftarrow x}p(R|t,\circ,x).}_{\text{people leaving from Nordics to } \circ}
\end{aligned}$$

We don't know the number of people arriving to Nordics from the outside world. However, we have an estimate of the total number of arriving infectious individuals, which we denote with $I^{\text{inflow}}$. We thus assume that the number of arriving Susceptible and Removed individuals is equal to the corresponding number of people leaving:

$$\begin{aligned}
D^{\text{long}}_{t,x\leftarrow\circ}p(I|t,x,\circ) &= I^{\text{inflow}}_{t,x};\\
D^{\text{long}}_{t,x\leftarrow\circ}p(S|t,x,\circ) &= D^{\text{long}}_{t,\circ\leftarrow x}p(S|t,\circ,x);\\
D^{\text{long}}_{t,x\leftarrow\circ}p(R|t,x,\circ) &= D^{\text{long}}_{t,\circ\leftarrow x}p(R|t,\circ,x);\\
p(I|t,\circ,x) &= I_{t,x}/N_{t,x}.
\end{aligned}$$

With these assumptions, model can then be written as

$$\begin{aligned}
S_{t+1,x} &= S_{t,\bullet}M^{\text{long}}_{t,x\leftarrow\bullet} - r_{t,x}S_{t,x};\\
I_{t+1,x} &= I_{t,\bullet}M^{\text{long}}_{t,x\leftarrow\bullet} + r_{t,x}S_{t,x} + I^{\text{inflow}}_{t,x} - I_{t,x}M^{\text{long}}_{t,\circ\leftarrow x} - \gamma I_{t,x};\\
R_{t+1,x} &= R_{t,\bullet}M^{\text{long}}_{t,x\leftarrow\bullet} + \gamma I_{tc}\\
r_{t,x} &= \beta_{t,x}I_{t,x}/N_{t,x},
\end{aligned}$$

where

$$M^{\text{long}}_{t,\circ\leftarrow x} = \frac{D^{\text{long}}_{t,\circ\leftarrow x}}{N_{t,x}}$$

In this model the local population sizes $N_x$ evolve over time as:

$$N_{t+1,x} = N_{t,\bullet}M^{\text{long}}_{t,x\leftarrow\bullet} + I^{\text{inflow}}_{t,x} - I_{t,x}M^{\text{long}}_{t,\circ\leftarrow x},$$

which means that they depend on the $\beta$ parameters. The figures below show the total population changes due to infections arriving ($I^{\text{inflow}}$) and pre-modelling estimated infected leaving, assuming $I_{t,x} \equiv 0.01N_{0,x}$. We see that the population changes remain $\leq 3\%$.

This version of the model can be visualized with the following flow diagram:

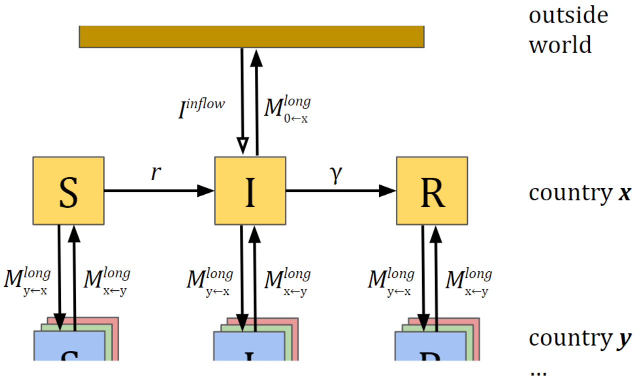


#### Adding short-term travellers between Nordic countries

We next let a series of mobility matrices $D^{\text{short}}_{t,d\leftarrow s}$ represent the number of individuals from the source country $s$ travelling to the destination country $d$ on day $t$ for a short-term visit (commuting). We assume here that both the origin and destination (target) conutries are in the four Nordics countries. We then define the relative mobility matrix for the short-term travels as

$$M^{\text{short}}_{t,d\leftarrow s} = \begin{cases} \nu \dfrac{D^{\text{short}}_{t,d\leftarrow s}}{N_{t,s}}, & \text{for } s \neq d; \\ 1 - \nu \dfrac{\sum_{x\neq s} D^{\text{short}}_{t,x\leftarrow s}}{N_{t,s}}, & \text{for } s = d. \end{cases}$$

Here $\nu$ is the portion of the day that a short-term traveller stays in the destination country. We assume $\nu = 0.5$, i.e. commuters spend half of the day home and half of the day in the destination country. The total number of individuals present in country $x$ on day $t$, including these with a short-term visit from other countries, would then be equal be equal to

$$N^*_{t,x} = \underbrace{N_{t,x}}_{\text{residents}} + \underbrace{\sum_{s,s\neq c} M^{\text{short}}_{t,x\leftarrow s} N_{t,s}}_{\text{commuters arriving}} - \underbrace{N_{t,x} \sum_{d,d\neq x} M^{\text{short}}_{t,d\leftarrow x}}_{\text{commuters leaving}} = \sum_{s} N_{t,s} M^{\text{short}}_{t,x\leftarrow s} = N_{t,\bullet} M^{\text{short}}_{t,x\leftarrow\bullet}$$

Note that here the change in the population size caused by commuters is small and can be ignored ($N_{t,x} \approx N^*_{t,x}$). The figure below shows the fluctuations in the population size, caused by daily commuters. They do not exceed 0.04%.

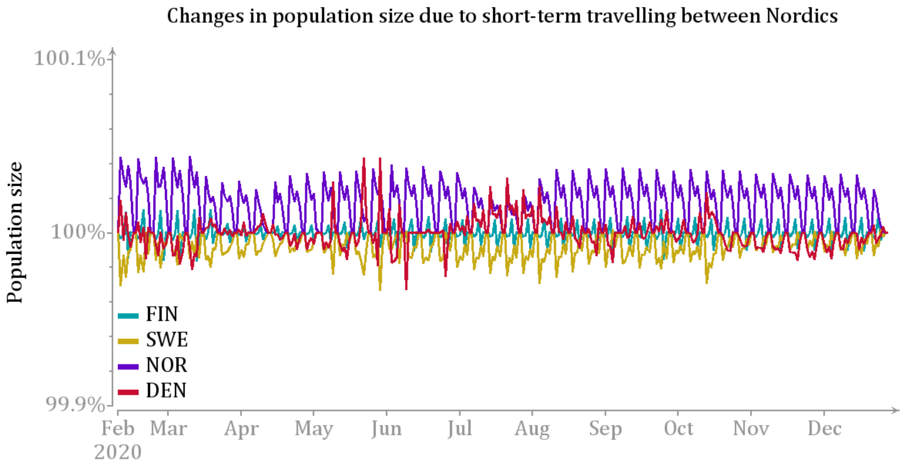


In analogy to long-term travel, we assume that the probabilities for the individuals to be susceptible, infectious or removed are equal to the proportional sizes of their compartments in the source country. The total number of infectious individuals (including commuters) present in country $x$ on day $t$ is then be equal to

$$I^*_{t,x} = I_{t\bullet} M^{\text{short}}_{t,x\leftarrow\bullet},$$

and the risk of being infected in country $x$ equals

$$r_{t,x} = \beta_{t,x} I^*_{t,x} / N_{t,x} = \beta_{t,x} I_{t,\bullet} M^{\text{short}}_{t,x\leftarrow\bullet} / N_{t,x}.$$

Among the susceptibles of country $x$, $M^{\text{short}}_{t,x\leftarrow x}$ we be exposed to the infection risk in the home country and $M^{\text{short}}_{t,y\leftarrow x}$ in the country $y$. Therefore the final equation reads

$$\begin{aligned} S_{t+1,x} &= S_{t,\bullet} M^{\text{long}}_{t,x\leftarrow\bullet} - r_{t,\bullet} M^{\text{short}}_{t,\bullet\leftarrow x} S_{t,x}; \\ I_{t+1,x} &= I_{t,\bullet} M^{\text{long}}_{t,x\leftarrow\bullet} + r_{t,\bullet} M^{\text{short}}_{t,\bullet\leftarrow x} S_{t,x} + I^{\text{inflow}}_{t,x} - I_{t,x} M^{\text{long}}_{t,\circ\leftarrow x} - \gamma I_{t,x}; \\ R_{t+1,x} &= R_{t,\bullet} M^{\text{long}}_{t,x\leftarrow\bullet} + \gamma I_{t,c}; \\ r_{t,x} &= \beta_{t,x} I_{t\bullet} M^{\text{short}}_{t,x\leftarrow\bullet} / N_{t,x}. \end{aligned}$$

We do not model short-term visits between Nordic countries and the outside world, because such flows are expected to be vanishigly small during the pandemic. Finally, the full model can be visualized with the following flow diagram:

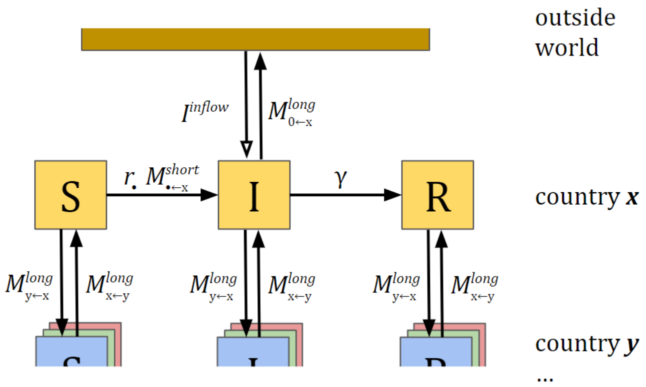
outside world
$I^{inflow}$
$M^{long}_{0\leftarrow x}$
S
$r_{\bullet}\ M^{short}_{\bullet\leftarrow x}$
I
γ
R
country ***x***
$M^{long}_{y\leftarrow x}$
$M^{long}_{x\leftarrow y}$
$M^{long}_{y\leftarrow x}$
$M^{long}_{x\leftarrow y}$
$M^{long}_{y\leftarrow x}$
$M^{long}_{x\leftarrow y}$
country ***y***
...
