## Appendix S3 for "The influence of cross-border mobility on the COVID-19 epidemic in Nordic countries"

### The influence of cross-border mobility on the COVID-19 epidemic in Nordic countries
### S3 Appendix: Additional Results

November 2, 2023

Shubin Mikhail[1 *], Hilde Kjelgaard Brustad[2], Jørgen Eriksson Midtbø[3], Felix Günther[4], Laura Alessandretti[5], Tapio Ala-Nissila[6, 7], Gianpaolo Scalia Tomba[4, 8], Mikko Kivelä[9], Louis Yat Hin Chan[3], Lasse Leskelä[1]

**1** Department of Mathematics and Systems Analysis, Aalto University, Espoo, Finland
**2** Oslo Center for Biostatistics and Epidemiology, Oslo University Hospital, Norway
**3** Department of Method Development and Analytics, Norwegian Institute of Public Health, Oslo, Norway
**4** Department of Mathematics, Stockholm University, Sweden
**5** DTU Compute, Technical University of Denmark, Copenhagen, Denmark
**6** Quantum Technology Finland Center of Excellence, Department of Applied Physics, Aalto University
**7** Interdisciplinary Centre for Mathematical Modelling and Department of Mathematical Sciences, Loughborough University, United Kingdom
**8** Department of Mathematics, University of Rome Tor Vergata, Italy
**9** Department of Computer Science, Aalto University, Espoo, Finland

*

In this appendix, we present some additional results complementing those in the main text.

#### 1 Model estimates

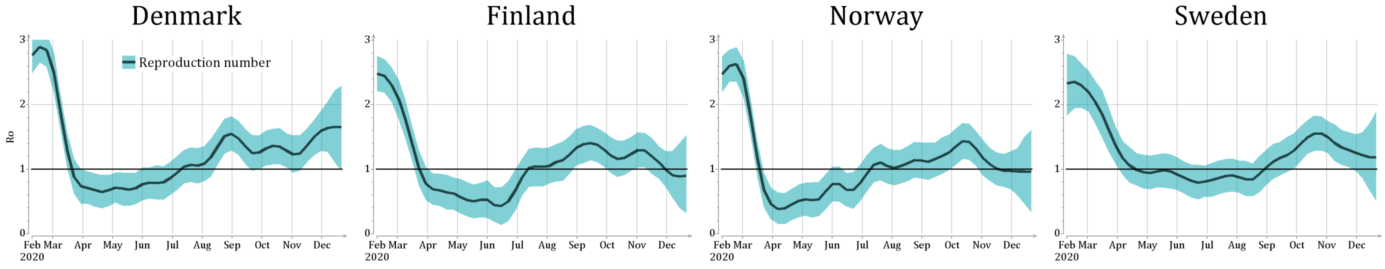


Figure 1: Estimates of the reproduction number $\mathcal{R}_{t,x}$. Posterior mean and 90% credible intervals.

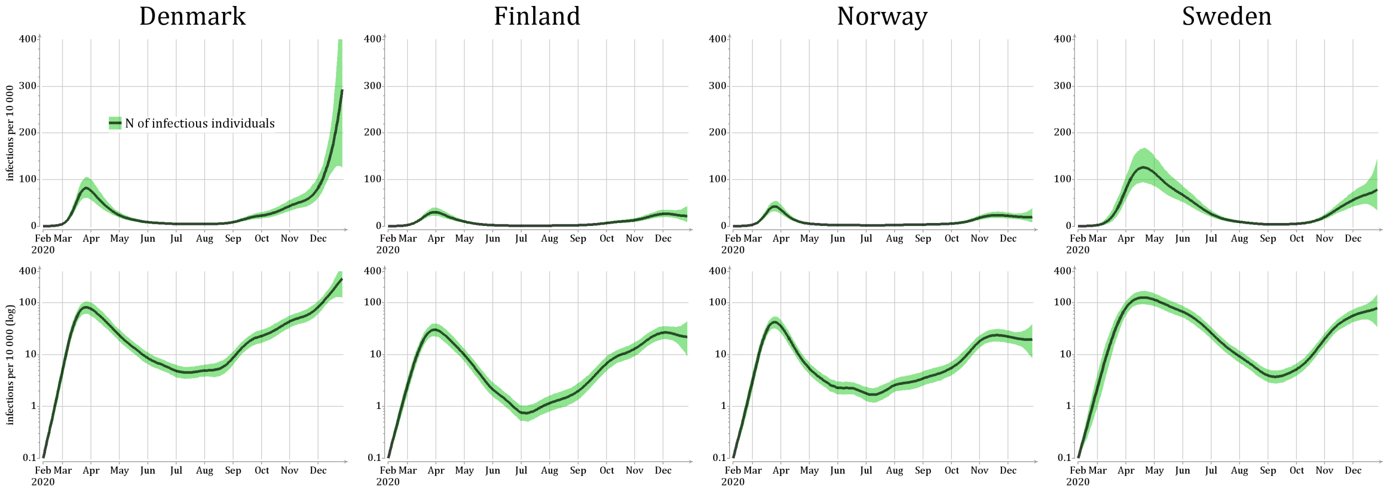


Figure 2: Estimated numbers of infectious individuals $I_{t,x}$, in linear and log scales. Posterior mean and 90% credible intervals.

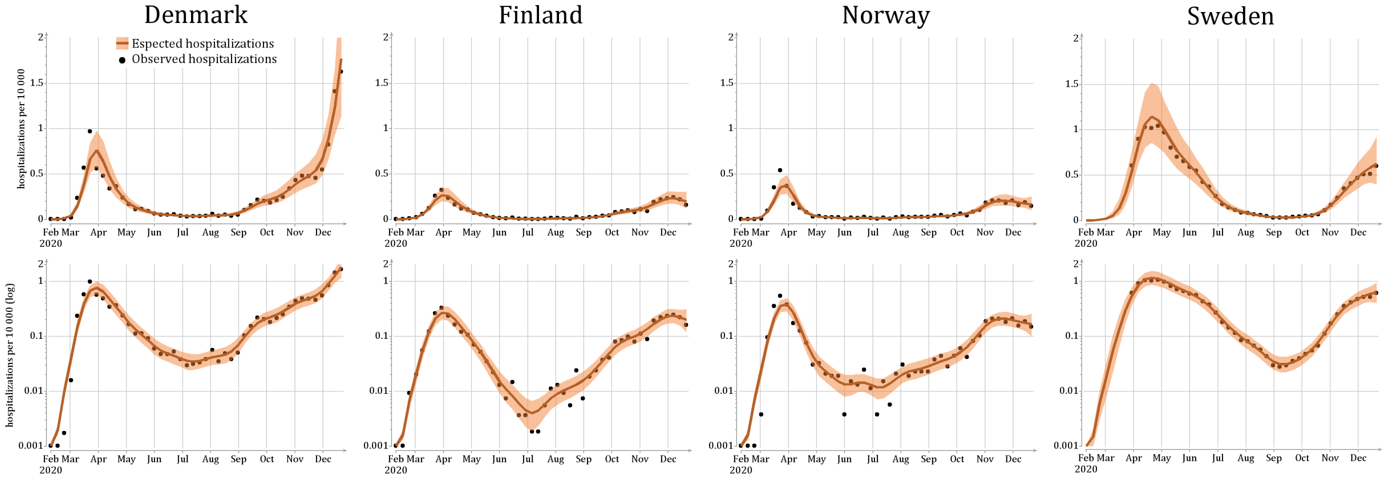


Figure 3: The expected number of hospitalizations per week $E_{w,x}$ and the corresponding observed numbers observed numbers $H_{w,x}$ in linear and log scales. Posterior mean and 90% credible intervals, dots present the observed data.

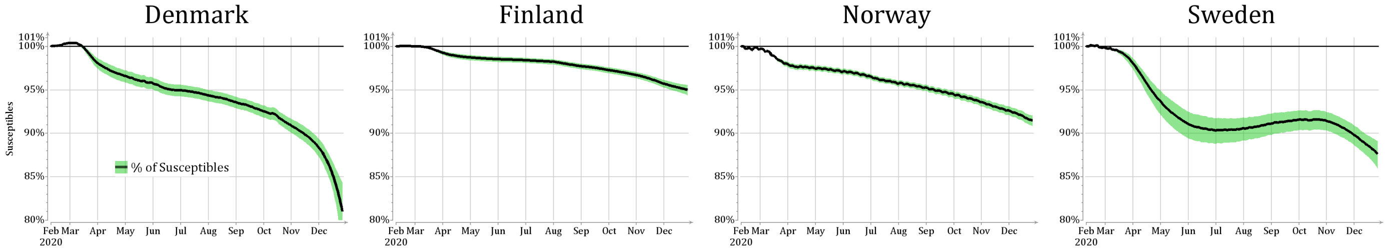


Figure 4: Estimated portion of Susceptibles in the population $S_{t,x}/N_{0,x}$. Posterior mean and 90% credible intervals.

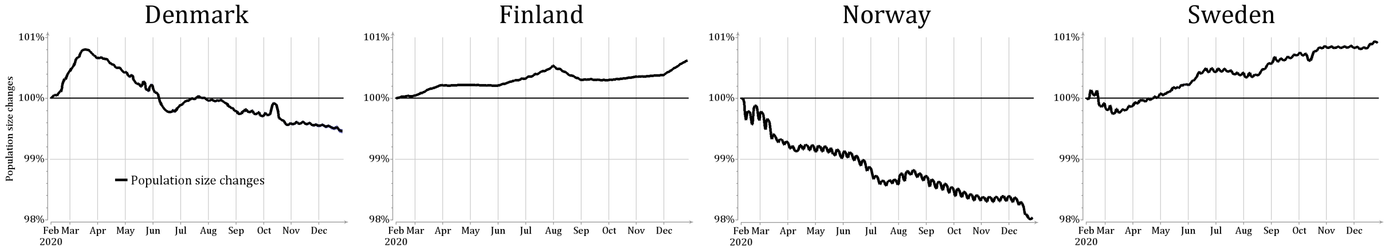


Figure 5: Changes in the population size in countries due to mobility $N_{t,x}/N_{0,x}$, relative to the starting population size. Posterior mean and 90% credible intervals.

### 2 Primary effect estimations

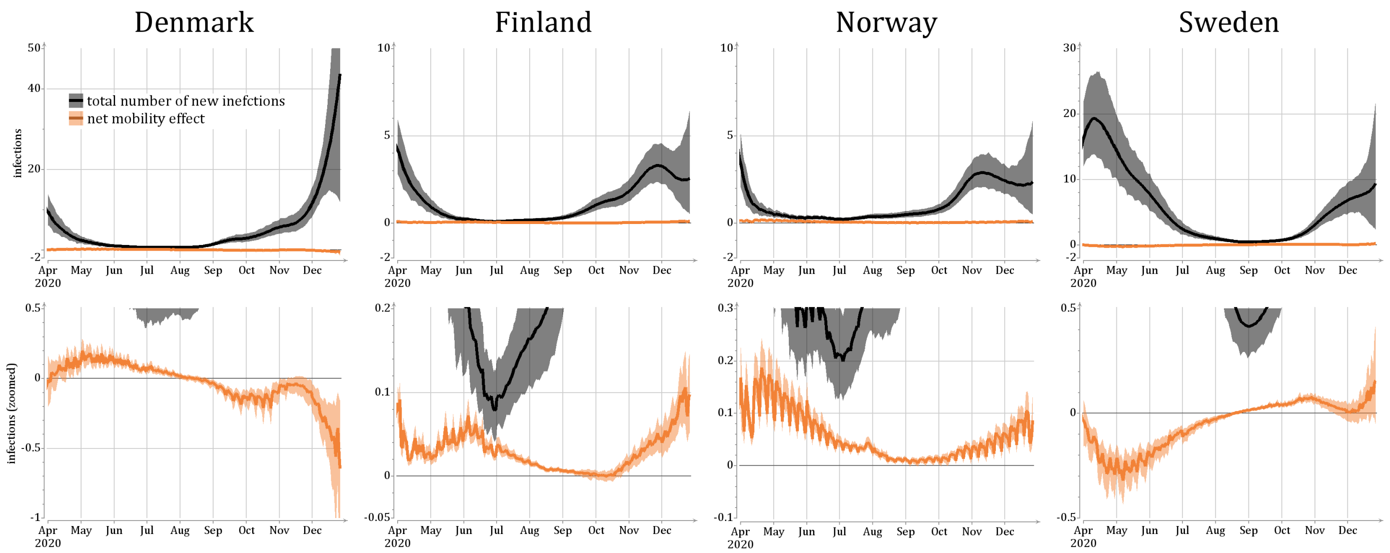


Figure 6: Estimates of the total number of new infections per day $i_{t,x}^{\text{total new}}$ local and the net mobility effect $Q_{t,x}$, shown at different linear scales.

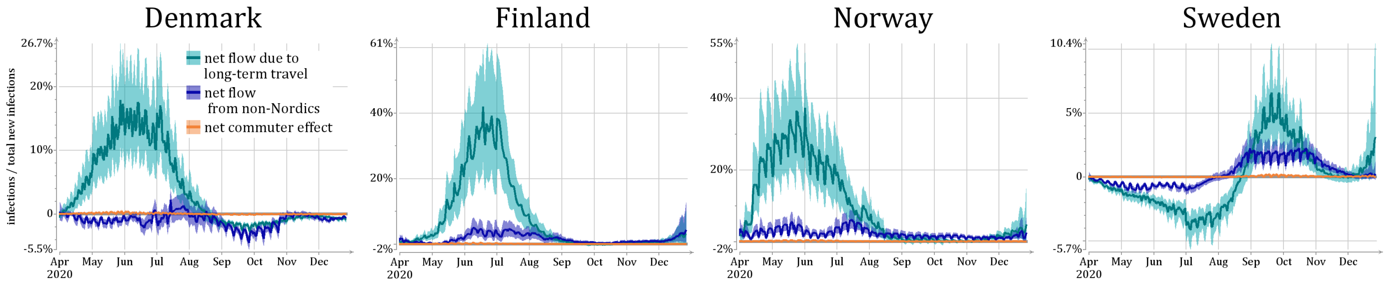


Figure 7: Estimates of new non-local infection by source, divided by the total number of new infection: $Q_{t,x}^{\text{long term}}/i_{t,x}^{\text{total new}}$, $Q_{t,x}^{\text{non-Nordic}}/i_{t,x}^{\text{total new}}$ and $Q_{t,x}^{\text{commuters}}/i_{t,x}^{\text{total new}}$. Posterior mean and 90% credible intervals.

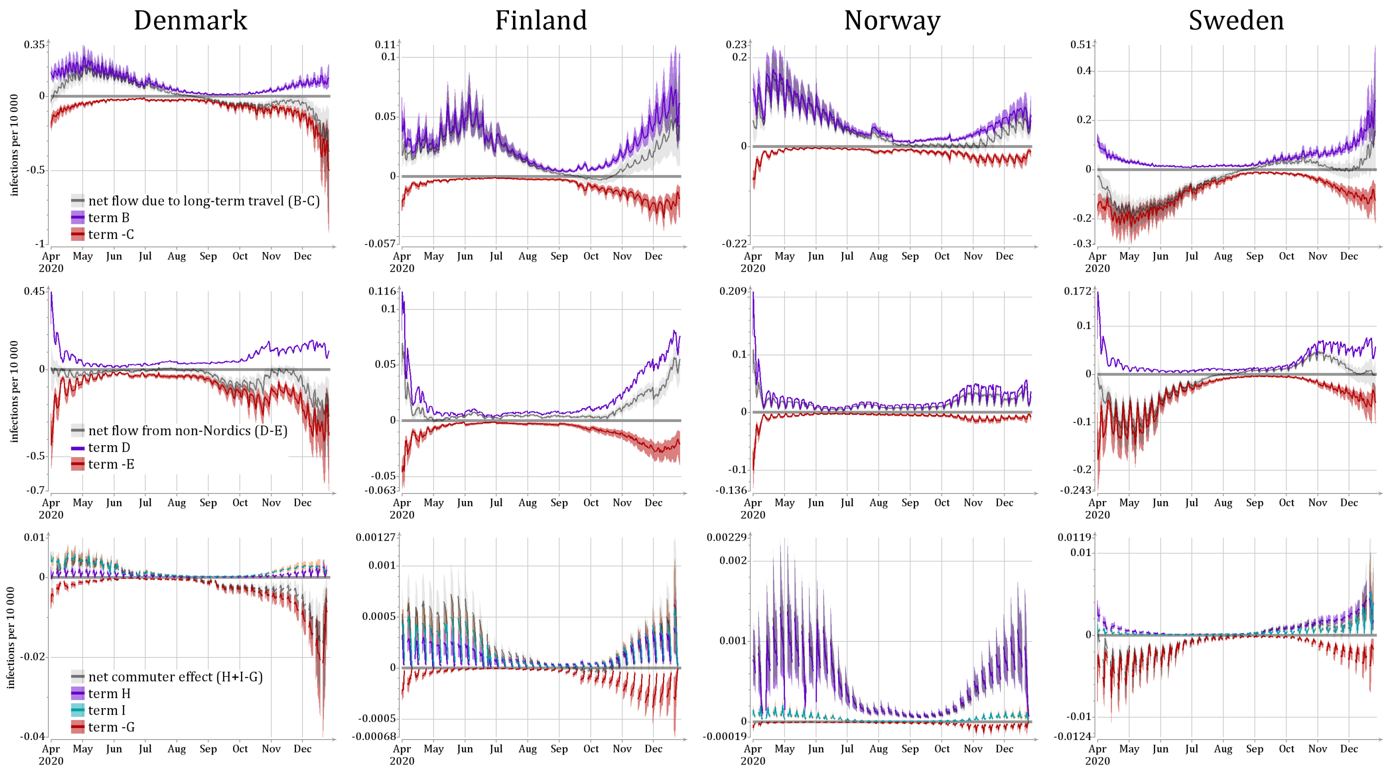


Figure 8: Estimates of new non-local infection by source, divided by the total number of new infection. Posterior mean and 90% credible intervals.

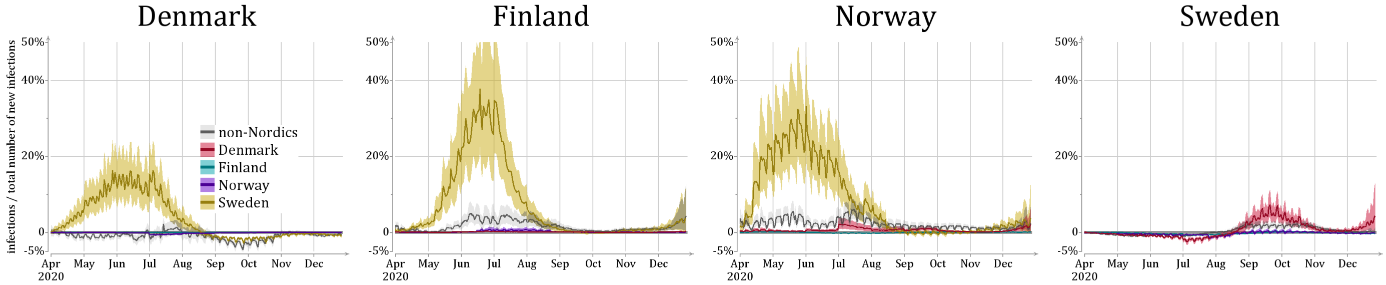


Figure 9: Estimates of non-local infections by country per day, divided by the total number of new infection.. Lines show posterior mean and colored areas show 90% posterior intervals.

### 3 Secondary effect estimations

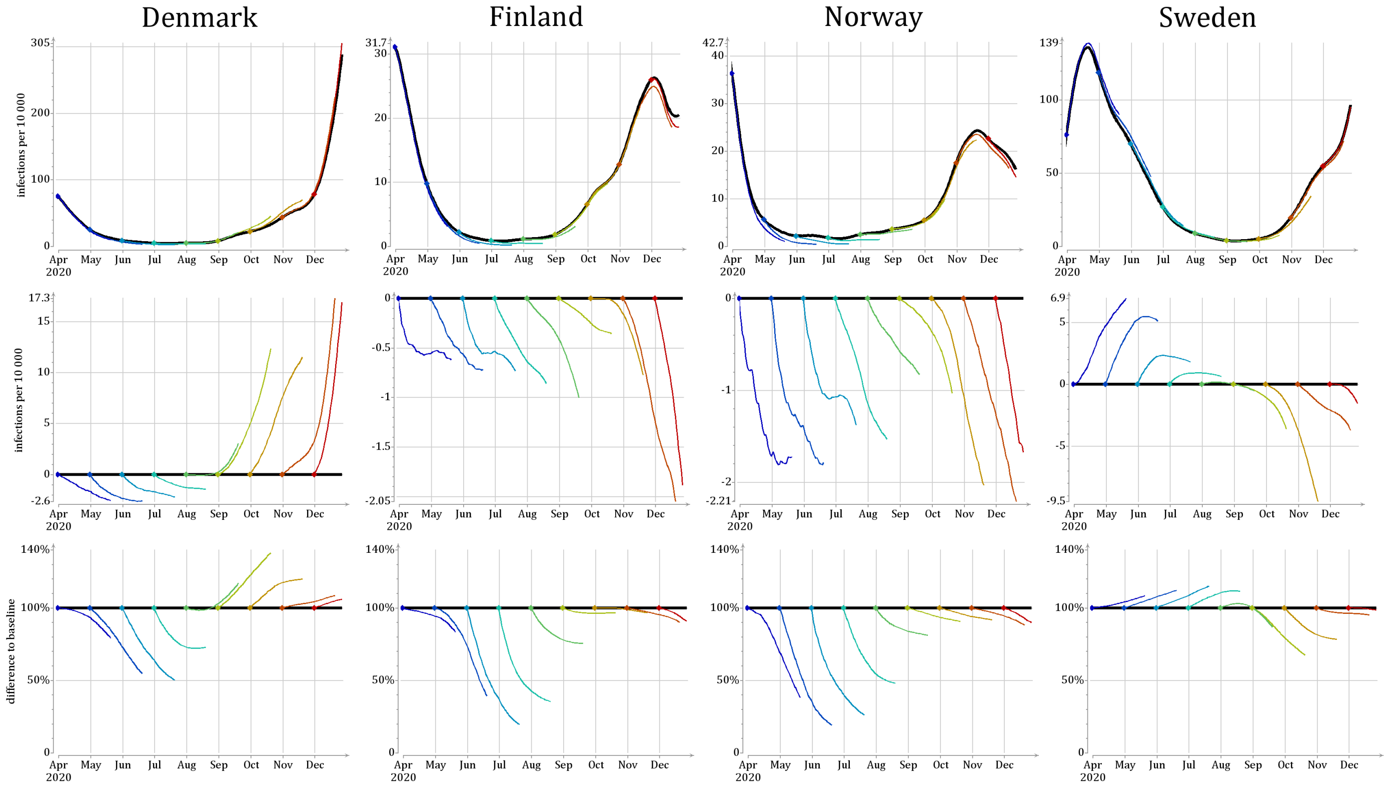


Figure 10: Alternative representation of the counterfactual analysis. Top row: using linear scale. Mid row: as a difference between counterfactual and baseline scenario. Bottom row: counterfactual divided by baseline scenario.

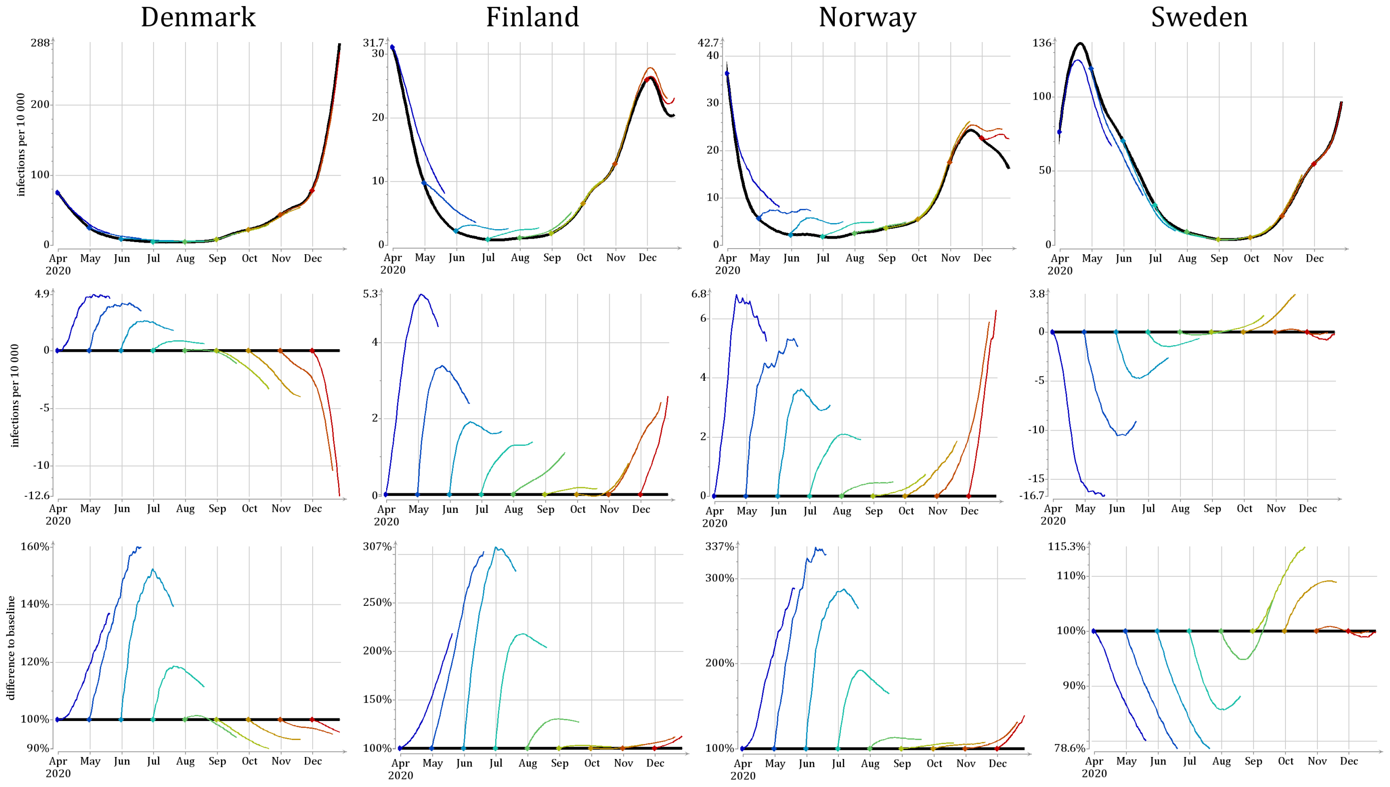


Figure 11: Alternative representation of the counterfactual analysis. Top row: using linear scale. Mid row: as a difference between counterfactual and baseline scenario. Bottom row: counterfactual divided by baseline scenario.
